# Pathway-specific polygenic risk scores reveal underlying mechanisms of cerebral small vessel disease

**DOI:** 10.64898/2026.09.10.26360585

**Authors:** Tim D’Aoust, Ami Tsuchida, Carole Dufouil, Catherine Helmer, Philippe Amouyel, Jean-Charles Lambert, Quentin Le Grand, Stéphanie Debette

## Abstract

White matter hyperintensities (WMH), a key MRI-marker of cerebral small vessel disease (cSVD), are common in older adults and associated with an increased risk of stroke and dementia. The latest WMH genome-wide association study (GWAS) identified 27 loci involving genes enriched for extracellular matrix, myelination, and membrane transport. Genetically predicted WMH correlates with white matter microstructural alterations in young adults and shows causal effects on stroke and dementia. However, biological pathways underlying WMH and their contribution to clinical outcomes remain unclear, and validated polygenic risk scores (PRS) for WMH are lacking. We applied global and pathway-specific PRS (ps-PRS) to generate robust WMH-PRS and identify biological pathways contributing to WMH across the lifespan.

We leveraged the largest European-ancestry WMH-GWAS (*N=46,944*) and data from 15,320 UK Biobank participants with MRI (UKB-MRI) (mean age=67±7.7 years) to construct candidate WMH global-PRS and 3,794 ps-PRS based on canonical pathways from the Molecular Signatures Database (v2023.2). The best WMH global-PRS in UKB-MRI (*P=1.78×10^-193^*; ΔR^2^=+4.1%) predicted WMH in independent cohorts: young adults (i-Share, *N=1,578*, age=22±2.3; *P=0.0024*; ΔR^2^=+0.5%), older community-dwellers (Three City-Dijon, *N=1,443*, age=73±4.1; *P=1.38×10^-9^*; ΔR^2^=+2.2%), and memory-clinic patients (Memento, *N=1,831*, age=71±8.5; *P=1.11×10^-19^*; ΔR^2^=+3.3%). In UK Biobank (*N* up to 355,180), WMH global-PRS was associated with incident stroke (HR=1.06 [1.038, 1.082], *P=2.6×10^-8^*), including both ischemic stroke (HR=1.061[1.037, 1.086], *P=1.6×10^-8^*) and intracerebral hemorrhage (HR=1.083[1.026, 1.143], *P=0.004*), as well as with incident all-cause dementia (HR=1.051 [1.030, 1.080], *P=1×10^-5^*) and its vascular or mixed sub-type (HR=1.168[1.103, 1.237], *P=1.1×10^-7^*).

Permutation-based pathway enrichment, performed in UKB-MRI, identified 127 ps-PRS consistently enriched for WMH and clustering into ten biological domains. Of these, 61 ps-PRS (six clusters and 55 individual pathways) were enriched in at least one follow-up cohort: 14 in older community-persons, 17 in young adults, and 37 in memory-clinic patients.

Secondary analyses highlighted four ps-PRS involved in lipid metabolism, ciliogenesis, and signal transduction enriched in both young and older adults and associated with stroke and dementia. In the memory-clinic some ps-PRS, notably involved in sphingolipid metabolism, were also associated with dementia. Seven ps-PRS, mostly lipid-related, showed evidence of modulation by hypertension.

In summary, we generated a validated WMH global-PRS showing robust association with cSVD clinical complications and introduce a multi-cohort WMH ps-PRS framework that reveals candidate biological pathways with differential associations across the lifespan and clinical outcomes. These findings may inform precision prevention and drug development for cSVD.

## Introduction

Cerebral small vessel disease (cSVD), characterized by pathological changes in the structure and function of small brain vessels, is a leading cause of ischemic stroke (IS) and intracerebral hemorrhage (ICH), and the main pathological substrate underlying the vascular contribution to cognitive decline and dementia^1^. Most often, cSVD manifests as ‘covert’ vascular lesions, detectable on brain magnetic resonance imaging (MRI) in the absence of clinical symptoms. Covert cSVD is extremely prevalent in older adults and is associated with increased risk of stroke, dementia, depression, and mortality, representing a major target to prevent age-related brain diseases in the general population^2^. Cardiovascular risk factors, particularly hypertension, are associated with cSVD, but only account for approximately 2% of the variance in cSVD lesion burden^3^, suggesting a substantial role of genetic and other factors. While optimal management of cardiovascular risk factors can slow down the progression of cSVD^2^, there are currently no mechanism-based treatments targeting the disease process.

The most studied cSVD MRI-marker is white matter hyperintensities of presumed vascular origin (WMH)^4^. Histologically, they are heterogenous, representing varying degrees of ischemic demyelination, axonal loss, and gliosis^5^. Recent developments now enable precise quantification of WMH volume using automated algorithms in large-scale population-based cohorts, even in those with very small WMH volumes (e.g. young adults)^6^. WMH volume is associated with cSVD-related complications, primarily stroke, dementia^4^, depression^7–9^, and cognitive performance^10,11^.

Genome-wide association studies (GWAS) of cSVD MRI-markers have begun to unravel molecular mechanisms underlying cSVD identifying 27 genetic loci associated with WMH volume^12–15^, with an overrepresentation of genes involved in extracellular matrix and cell membrane structure, myelination, and membrane transport^12^. Polygenic risk scores (PRS) derived from GWAS aggregate the effects of numerous independent risk variants across the genome to capture global genetic risk for complex traits, and have demonstrated ability to identify individuals at high genetic risk of multifactorial diseases^16^. Interestingly, PRS focusing on genome-wide significant risk variants of WMH (also referred to as weighted genetic risk score, GRS), have been linked to additional cSVD MRI-markers of white matter microstructure, including diffusion MRI metrics in young adults, suggesting early genetic influences on disease pathogenesis^13,17^. However, the ability of these PRS to predict clinical cSVD complications has not been thoroughly explored.

Furthermore, besides PRS representing ‘global’ genomic risk (global-PRS), aggregating genetic risk according to defined gene sets may allow to better characterize the biological pathways involved in cSVD across the lifespan^18^. Such pathway-specific PRS (ps-PRS) partition genetic liability into biologically meaningful pathways. Ps-PRS can be defined with *a priori* knowledge of pathological processes (e.g. ps-PRS for Aβ clearance or cholesterol metabolism^19^) or agnostically^20,21^, using data-driven approaches with pathway enrichment tools like MAGMA^22^ or canonical databases to define viable pathways^23–26^. This hypothesis-free approach has uncovered enriched pathways involved in neurological conditions such as Parkinson’s disease^21^, multiple sclerosis^27^, or AD-related changes in brain structure^20^. The advantage of ps-PRS over traditional pathway enrichment methods lies in its ability to obtain an estimate of an individual’s susceptibility to specific disease-related pathways. Associations of enriched pathways with disease complications or biomarkers can then be tested across independent cohorts representing different life stages and clinical contexts. For cSVD, ps-PRS approaches could uncover novel biological pathways and provide important context for the development of biomarkers and therapeutic targets across the age and disease spectrum.

Leveraging biobank-scale data and a large GWAS meta-analysis of WMH in older adults, we first optimized a WMH global-PRS to quantify overall genetic liability. Second, we applied an agnostic WMH ps-PRS approach to identify canonical pathways implicated in cSVD etiology. To capture the evolution of WMH determinants across the age and clinical spectrum, we then performed follow-up analyses of both WMH global-PRS and ps-PRS across diverse cohorts, in young adults, community-dwelling older adults, and memory-clinic patients. In secondary analyses we tested the association of the most robust ps-PRS with incident stroke and dementia outcomes in the general population and memory-clinic settings, and probed modulation of WMH (ps-)PRS by hypertension.

## Materials and methods

An overview of our study design is provided in **Figure 1**.

**Figure 1.**
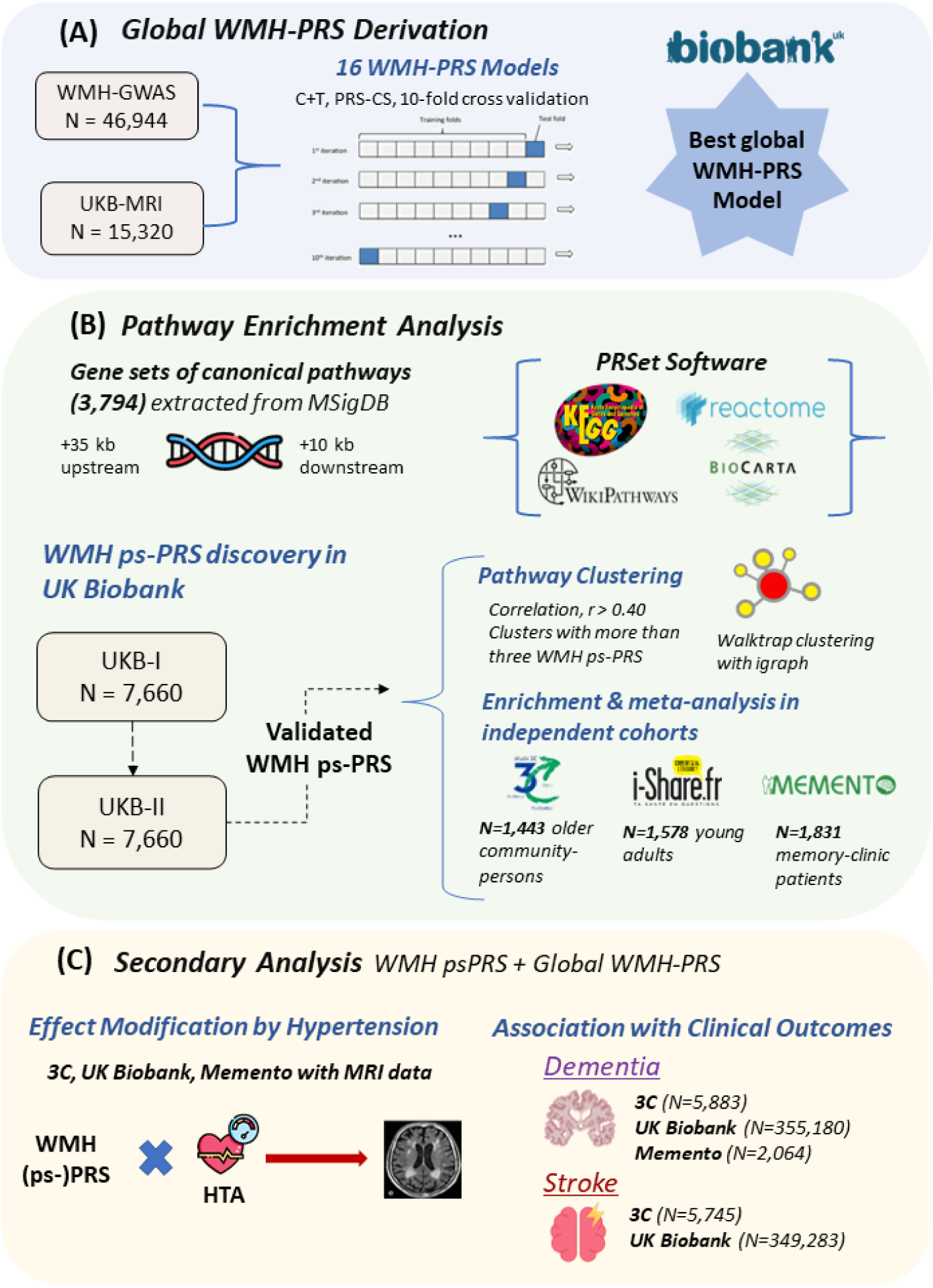
Overall study design. (A) Workflow used to derive and optimize WMH global-PRS using UK Biobank MRI data and 10-fold cross-validation. (B) Procedure to identify enriched WMH-psPRS and perform follow-up analyses in independent cohorts. (C) Secondary analysis was used to further investigate clinical and lifespan effects of selected pathways. MSigDB = molecular signature database; UKB = UK Biobank; C+T = clumping & thresholding. Kb = kilobases; 3C = Three Cities study; HTA = hypertension.

### Study populations

We leveraged multiple independent cohorts representing community and clinic-based populations across the lifespan. We excluded participants with pre-existing neurological conditions (e.g., brain tumors, Alzheimer’s Disease, Parkinson’s Disease, stroke, and multiple sclerosis), missing information on WMH volume or genetic data, and individuals of non-European ancestry (see **Supplementary Methods** for details on exclusion criteria in each cohort).

The UK Biobank is a prospective, population-based cohort of over 500,000 individuals with genetic data recruited between the ages of 40 and 69 between 2006 and 2010, and followed up longitudinally, with a large imaging sub-study (up to 100,000 participants). In our analysis of WMH volume, after exclusions, we focused on a subset of 15,320 participants (mean-age=67.1±7.2 years, 53.2% women) recorded after January 26th, 2020, to avoid overlap with participants included in the WMH GWAS meta-analysis used to derive the PRS^13^. Data from these participants (referred to as UKB-MRI henceforth) were used to optimize genome-wide WMH PRS and identify a set of validated ps-PRS representing canonical biological pathways enriched for WMH.

The Three Cities (3C) study is a population-based cohort comprising community-dwelling persons aged ≥65 years from three French cities, Dijon, Montpellier, and Bordeaux, recruited in 1999-2001 and followed every 2-3 years over 12-17 years^28^. In Dijon, participants aged <80 years and enrolled between June 1999 and September 2020 were invited to undergo brain MRI (N=1,924)^29^. After exclusions, our sample comprised 1,443 eligible participants (mean age=72.7±4.1 years, 60.2% women).

The internet-based Students Health Research Enterprise (i-Share) is a French prospective population-based study of university students aged 18-35 years^30^, in which a sub-set of participants were invited for brain MRI and genome-wide genotyping through the MRi-Share and bio-Share ancillary studies^31,32^. After exclusions, our sample comprised 1,578 eligible participants (mean age=22.1±2.3 years, 71.7% women).

The Memento study is a memory clinic-based cohort^33^ including 2,323 participants with MCI or subjective cognitive complaints recruited from 26 French memory clinics between 2011-2014 and followed up every 6-12 months over 5 years. After exclusions, 1,831 participants were included in the present analysis (mean-age=71.1±8.5 years, 62.2% women).

### MRI Acquisition and WMH Derivation

Brain MRI scans were obtained with a 1.5 Tesla (3C: Siemens Magnetom) or 3 Tesla (UK Biobank: Siemens Skyra; i-Share: Siemens Prisma) scanner using standardized protocols, as detailed elsewhere^29,32,34,35^. For Memento, 1.5 or 3 Tesla scans were collected at memory clinics and harmonized by the Center for Automated Treatment of Images (CATI; cati-neuroimageing.com)^36^. A fully automated software to quantify WMH volume from T1-weighted and FLAIR images was implemented in i-Share, Memento, and UK Biobank^6,37,38^. Uniquely in 3C-Dijon, T1- and T2-weighted and proton density images were used for WMH segmentation^39^. Total WMH volume was rank-based inverse-normal transformed to normalize and standardize the trait and allow for better comparison across studies. In the i-Share cohort, due to the large number of equally ranked values, we used indirect inverse-normal transformation of WMH volume, which involves applying inverse-normal transformation on WMH-residuals after adjustment for sex, age at MRI, estimated total intracranial volume, and the first four principal components of population stratification (PCs). More detailed description of MRI acquisitions and WMH derivation in each study is available in the **Supplemental Material.**

### Genotyping, quality control, and imputation

Genome-wide genotyping was performed using the Affymetrix UK BiLEVE Axiom Array and UK Biobank Axiom Array in UK Biobank, Affymetrix Axiom Precision Medicine Research Array in i-Share, the Illumina Human610Iquad BeadChips in 3C-Dijon, and the Illumina GSA in Memento. Quality control and imputation procedures are described elsewhere^31,40^ and in the **Supplementary Material**.

### WMH Polygenic Risk Score Derivation

We used the largest WMH GWAS summary statistics comprising 46,944 older adults of European-ancestry to derive both WMH global-PRS and ps-PRS. These summary statistics were re-generated from the original study excluding 3C-Dijon to avoid sample overlap^13^. In addition, we removed all single nucleotide polymorphisms (SNPs) that were duplicated, multi-allelic, with minor allele frequency <1%, not available or mismatched in the 1000 Genomes European reference panel (and across cohorts)^41^, or with poor imputation quality (imputation score <0.8).

#### WMH global-PRS

To first optimize WMH global-PRS, we derived 16 candidate WMH PRS models in 15,320 UK Biobank participants using different parameterizations for two established PRS methods: ten models based on ‘clumping and thresholding’^42^ with varying p-value thresholds, and six based on PRS-CS^43^ (**Figure 1A**, PRS methods description in **Supplementary Methods**). PRS models were calculated using the GenoPred pipeline which provides a streamlined, standardized framework to perform quality control and derive PRS in target datasets^44^. The 1000 Genomes European sample was used as an external linkage disequilibrium (LD) reference for both methods^41^.

We implemented 10-fold cross validation using the *caret* R package^45^ in order to select the WMH global-PRS that best predicts WMH volume in UK Biobank. All models for testing association with inverse-normal transformed WMH volume (field 25781, instance 2) were adjusted for age at MRI (field 21003, instance 2), age at MRI-squared, sex (field 31), total intracranial volume (field 26521), the first 10 PCs of population stratification (field 22009), and genotyping chip (field 22000). The best WMH-PRS was selected based on minimization of the root-mean-squared-error and used as the WMH global-PRS in all subsequent analyses. We then tested the association of the WMH global-PRS with WMH volume in the 3C-Dijon, i-Share, and Memento studies, followed by meta-analysis across cohorts using the inverse-variance weighted (IVW) method implemented in the *metafor* R package^46^.

#### WMH pathway-specific PRS

We aimed to identify a set of ps-PRS that were enriched for WMH in UK Biobank using the PRSet function implemented in PRSice-2 (v2.3.5)^42^. In an agnostic approach, we defined ps-PRS using gene sets from 3,794 curated biological pathways in the Canonical Pathway subset of Molecular Signatures Database (MSigDB v2023.2)^47^, and weights from the latest WMH GWAS^13^. Pathway databases contributing to MSigDB v2023.2 included Reactome, BioCarta, Kyoto Encyclopedia of Genes and Genomes (KEGG), pathway interaction database, and WikiPathways. To validate enriched pathways, we split 15,320 UK Biobank participants randomly into two sub-samples, UKB-I and UKB-II (**Figure 1B**). We used a permutation-based empirical p-value (*P_emp_*)<0.05 in UKB-I to filter candidate pathways and validated these pathways in UKB-II to select ps-PRS enriched for WMH. More details on PRSet pathway enrichment are presented in the **Supplementary Material**. For brevity, pathway source databases—REACTOME, KEGG, KEGG Medicus, WikiPathways, Pathway Interaction Database, and BioCarta—are prefixed in pathway names as R, KG, KG MED, WP, PID, and BC, respectively.

We then performed clustering using the *igraph* R package^48,49^ to identify relevant clusters of correlated pathways. Specifically, Walktrap clustering^50^ was performed on a graph network built from an adjacency matrix of correlations (r>0.4) between WMH ps-PRS. For clusters of more than three ps-PRS, representing a sufficient number of non-identical pathways, we defined ‘cluster ps-PRS’ using expanded gene sets to include all genes within pathways in the same cluster. We inputted the gene sets of these cluster ps-PRS into g:Profiler^51^ to perform functional profiling based on molecular functions, biological processes, and cellular components from Gene Ontology^52,53^. We assigned relevant names to each cluster based on the most significant Gene Ontology annotations from the full list of genes in the cluster and confirmed enrichment of cluster ps-PRS in UKB-I and UKB-II with PRSet.

Next, we followed up pathway enrichment of individual and cluster WMH ps-PRS enriched in UK Biobank (*P_emp_<0.05*) in the 3C-Dijon, i-Share, and Memento studies. Linear regression was also used in each study to obtain direct effect estimates for association of WMH ps-PRS with WMH volume, which were then meta-analyzed across studies using IVW. We used Bonferroni multiple testing correction after determining the number of independent tests while accounting for correlation between WMH ps-PRS in UKB-MRI using the matSPDlite script in R^54^. Linear regression analyses were performed using R version 4.1^55^. All pathway enrichment and linear regression analyses were adjusted for the same covariates as WMH global-PRS derivation, and cohort-specific variables (e.g., memory-clinic center in Memento, genotyping chip in UK Biobank). PRS were mean-zero standardized, and all associations were reported per 1SD of (ps-)PRS.

### Secondary Analyses

For secondary analyses, we selected the WMH global-PRS, six replicating cluster ps-PRS and 55 individual ps-PRS enriched in at least one follow-up cohort, in order to assess modifying effects of hypertension and explore associations of ps-PRS with clinical complications of cSVD.

#### Effect of hypertension on the association of WMH ps-PRS with WMH volume

In the UK Biobank MRI sample (UKB-I and UKB-II), we also explored whether associations of WMH global- or ps-PRS with WMH volume are modulated by hypertension, the strongest risk factor for cSVD.

For this analysis, we included all participants from the initial UK Biobank MRI sample with blood pressure information (blood pressure levels and anti-hypertensive medication intake) at the time of MRI (*N=12,520*, 81.7% of main analytical sample). Hypertension was defined by systolic blood pressure ≥140 mmHg or diastolic blood pressure ≥90 mmHg or anti-hypertensive medication intake (**Supplemental Material**). We divided the sample into sub-groups based on hypertension status and estimated the association of WMH global-PRS or ps-PRS with WMH volume among hypertensives and non-hypertensives using linear regression models adjusted for the same original covariates. Heterogeneity between subgroup-specific effect estimates was formally tested using Cochran’s Q test. These analyses were performed using R version 4.1^55^, and the *metafor* package^46^ to test for heterogeneity.

#### Association of WMH (ps-)PRS with cSVD complications, stroke and dementia

To analyze the association of enriched WMH (ps-)PRS with incident stroke and dementia, we used all samples of European ancestry with genetic data available in UK Biobank (without brain MRI to avoid sample overlap), 3C, and Memento (for dementia only). We investigated associations of WMH (ps-)PRS with any stroke, ICH and IS, and with all-cause dementia (ACD), AD and vascular or mixed dementia (VMD). For analyses of any incident stroke cases, we removed all prevalent stroke cases, and for analyses of stroke sub-types (IS or ICH), we additionally removed those with ‘undefined’ stroke to avoid misclassification bias. Similar exclusions were applied to incident dementia analyses.

We assessed the associations of WMH (ps-)PRS with incident stroke and dementia using Cox proportional hazards models. We used age as a time scale, from age at baseline visit to end-of-follow-up, censoring at death, end of follow-up or event of other sub-type (for sub-type models). All models were adjusted for sex, genetic PCs, and other study-specific covariates (e.g., study centre). For association analyses with dementia, models were additionally adjusted for *APOE-ε2* and *APOE-ε4* dosages. We adapted models in each cohort to meet Cox proportional hazard model assumptions. Clinical outcome ascertainment and study-specific exclusions and modelling decisions are fully described in the **Supplemental Material**.

## Results

Baseline characteristics of UK Biobank, 3C-Dijon, Memento, and i-Share studies are shown in **Table 1**.

**Table 1.** Baseline characteristics of brain MRI studies. SD = standard deviation.

|  | UKB-I<br>(N=7660) | UKB-II<br>(N=7660) | 3C-Dijon<br>(N=1443) | iSHARE<br>(N=1578) | Memento<br>(N=1831) |
| --- | --- | --- | --- | --- | --- |
| **Age at MRI** |  |  |  |  |  |
| Mean (SD) | 67.2 (7.16) | 67.0 (7.17) | 72.7 (4.12) | 22.1 (2.26) | 71.1 (8.52) |
| Median [Min, Max] | 67.0 [52.0, 85.0] | 67.0 [51.0, 85.0] | 72.4 [65.2, 85.5] | 21.7 [18.1, 34.9] | 71.9 [32.9, 92.9] |
| **Sex** |  |  |  |  |  |
| Female | 4070 (53.1%) | 4077 (53.2%) | 869 (60.2%) | 1131 (71.7%) | 1138 (62.2%) |
| Male | 3590 (46.9%) | 3583 (46.8%) | 574 (39.8%) | 447 (28.3%) | 693 (37.8%) |
| **WMH Volume (cm3)** |  |  |  |  |  |
| Mean (SD) | 5.57 (7.23) | 5.65 (7.28) | 5.57 (4.93) | 1.31 (1.29) | 9.51 (12.6) |
| Median [Min, Max] | 3.17 [0.0990, 115] | 3.23 [0.0930, 94.6] | 4.07 [0.499, 50.0] | 1.03 [0, 15.7] | 5.44 [0.0800, 123] |
| **Systolic Blood Pressure** |  |  |  |  |  |
| Mean (SD) | 147 (21.1) | 148 (20.8) | 149 (22.8) | 124 (12.6) | 137 (18.1) |
| Missing | 1871 (24.4%) | 1882 (24.6%) | 0 (0%) | 713 (45.2%) | 74 (4.0%) |
| **Diastolic Blood Pressure** |  |  |  |  |  |
| Mean (SD) | 79.6 (11.2) | 79.9 (10.7) | 84.9 (11.6) | 76.1 (8.05) | 77.3 (10.1) |
| Missing | 1859 (24.3%) | 1875 (24.5%) | 0 (0%) | 713 (45.2%) | 74 (4.0%) |
| **Antihypertensive Medication** |  |  |  |  |  |
| Mean (SD) | 0.272 (0.445) | 0.262 (0.440) | 0.426 (0.495) | 0 (0) | 0.333 (0.471) |
| Missing | 95 (1.2%) | 107 (1.4%) | 0 (0%) | 713 (45.2%) | 74 (4.0%) |
| **Hypertension** |  |  |  |  |  |
| Non-Hypertensive | 1735 (22.7%) | 1679 (21.9%) | 626 (43.4%) | 840 (53.2%) | 1077 (58.8%) |
| Hypertensive | 4530 (59.1%) | 4576 (59.7%) | 817 (56.6%) | 25 (1.6%) | 680 (37.1%) |
| Missing | 1395 (18.2%) | 1405 (18.3%) | 0 (0%) | 713 (45.2%) | 74 (4.0%) |

### WMH global-PRS

In 15,320 UKB-MRI participants, the PRS-CS model (ϕ=auto) was best able to predict WMH volume in 10-fold cross-validation, and was strongly associated with WMH volume (Beta=0.204 95%CI [0.190, 0.217]; *P=1.78×10^-193^*, variance explained (R^2^) over base model with covariates only=+4.1%). The predictive ability of WMH global-PRS was sensitive to both PRS method and parameterization with R^2^ ranging from 1.8-4.1% over the base model with covariates only (Base model R^2^=26.9%, **Supplementary Table 1**). - We used the PRS-CS model (ϕ=auto) to represent WMH global-PRS in all subsequent analyses.

The WMH global-PRS was significantly associated to WMH volume in 3C-Dijon (*N=1,443*, Beta=0.153 [0.104, 0.202], *P=1.38×10^-9^*, R^2^= +2.2%), Memento (*N=1,831*, Beta=0.197 [0.155, 0.239], *P=1.11×10^-19^*, R^2^= +3.3%), and, to a lesser extent but significantly, in the much younger i-Share cohort (*N=1,578*, Beta=0.079 [0.028, 0.130], *P=0.0024*, R^2^=+0.5%). IVW meta-analysis across all four cohorts yielded a significant association between WMH global-PRS and WMH volume (Beta=0.161 [0.106, 0.216], *P=1.74×10^-8^*).

### WMH ps-PRS

In UKB-I (*n=7,660*), we identified 369 canonical pathways (out of 3,794) enriched for WMH (*P_emp_<0.05*). In UKB-II, 127 WMH of these 369 ps-PRS were also enriched and carried forward in pathway clustering and follow-up analyses in independent cohorts. All 127 WMH ps-PRS were enriched in the direction of increased WMH volume. Using Walktrap clustering on the 127 WMH ps-PRS we identified ten clusters of more than three ps-PRS encompassing 63/127 enriched pathways (49.6%) (Figure 2A, **Supplementary Table 2**). Full enrichment results for all 127 individual and 10 cluster ps-PRS in each cohort are included in **Supplementary Tables 3-6**.

**Figure 2.**
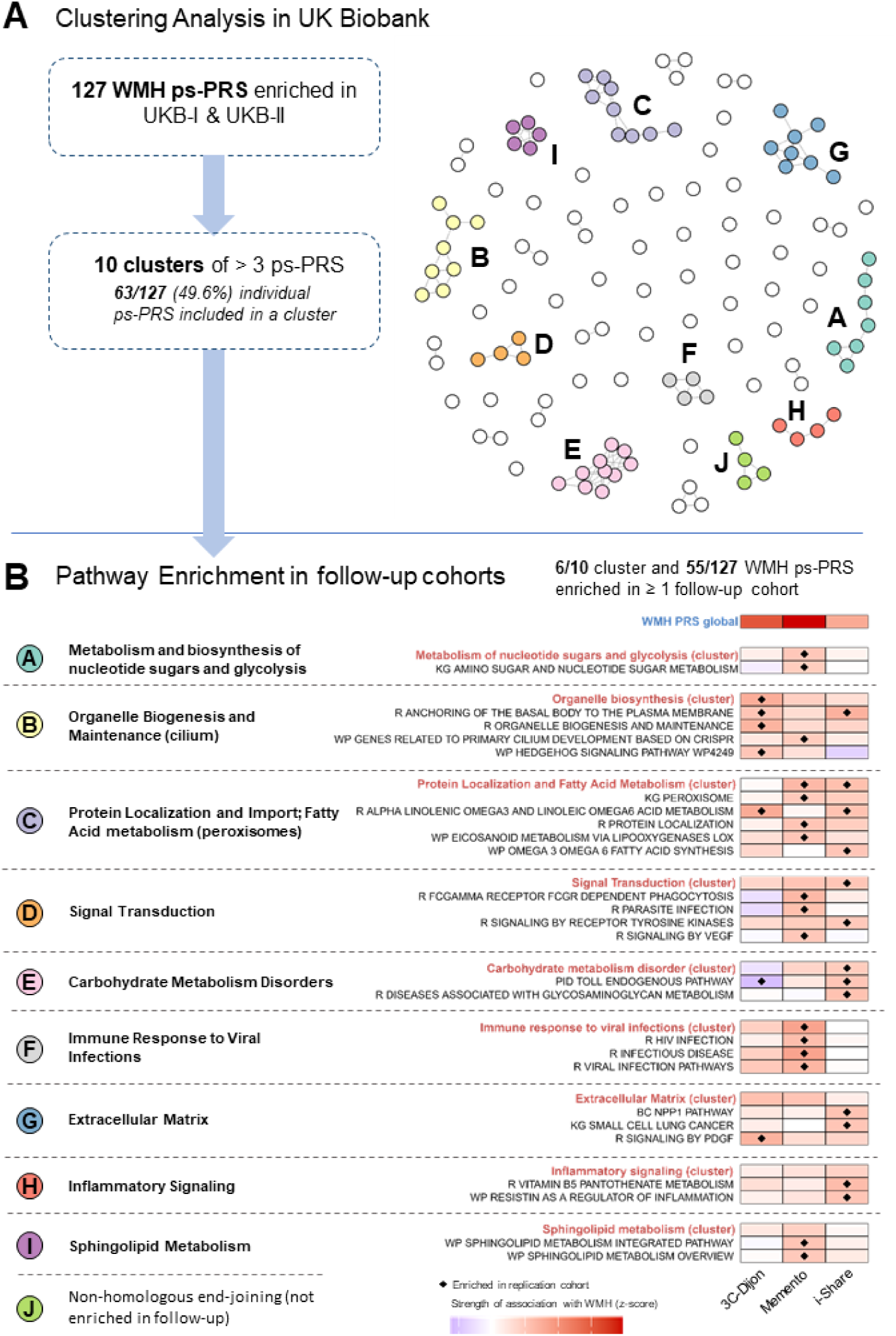
**(A)** Clustering of the 127 enriched WMH ps-PRS in UK Biobank. Clusters of more than 3 ps-PRS were defined based on Gene Ontology terms. Cluster-based ps-PRS were carried forward in follow-up analysis. **(B)** Pathway enrichment of 127 WMH ps-PRS were performed in independent follow-up cohorts (3C-Dijon, Memento, i-Share). Shown here are pathway enrichment and association with WMH volume in follow-up cohorts of replicating cluster-based WMH ps-PRS (in red) and individual WMH ps-PRS belonging to a cluster. Letter- and color-based indexing corresponds to the clusters in panel (A). WMH = white matter hyperintensities; ps-PRS = pathway-specific polygenic risk scores; R = Reactome; PID = Pathway Interaction Database; WP = Wiki Pathways; KG = KEGG.

No ps-PRS cluster was enriched in all three follow-up cohorts, but enrichment for six out of ten ps-PRS clusters was observed in at least one independent follow-up cohort (**Figure 2B**). The *protein localization and fatty acid metabolism* cluster (comprising 8 correlated pathways and 247 genes) was enriched in both young adults and memory-clinic patients. The *signal transduction* and *carbohydrate metabolism (disorders)* cluster (4 pathways, 623 genes) were enriched in young adults, the *organelle biogenesis* cluster (8 pathways, 415 genes) was enriched in older community-persons, and the *immune response to viral infections* (4 pathways, 1,019 genes) and *metabolism and biosynthesis of nucleotide sugars and glycolysis* (7 pathways, 111 genes) clusters were enriched in memory-clinic patients. Although the *extracellular matrix*, *sphingolipid metabolism,* and *inflammatory signalling* did not replicate at the cluster level, several individual ps-PRS included in these clusters did (**Figure 2B**).

At the individual ps-PRS level, 55/127 ps-PRS (of which 26 within and 29 outside clusters) were enriched in at least one follow-up cohort: 14 in older community-persons, 17 in young adults, and 37 in memory-clinic patients **(Figure 2B, Supplementary Fig. 1)**. Three pathways were enriched in both older community-persons and memory-clinic patients relating to *KG MED microtubule depolymerization* and two *R elastic fiber* pathways. Two pathways were enriched in both young and older community-dwelling adults: *R anchoring of the basal body to the plasma membrane* and *R alpha linolenic omega-3 and linoleic omega-6 metabolism.* One pathway, *PID ephrin type-A receptor 2 forward signaling,* was enriched in memory-clinic patients and young adults.

Finally, we estimated the associations of the 61 replicating WMH ps-PRS (six clusters and 55 individual ps-PRS) with WMH volume in each cohort using linear regression models adjusted for the same covariates and performed IVW meta-analysis across all cohorts (including UK Biobank). Using Bonferroni multiple testing correction (*P<0.00101*; 49 independent tests determined in UKB-MRI), 35 WMH ps-PRS were significantly associated to WMH volume in meta-analysis, showing consistent association across all cohorts (**Supplementary Tables 7-8**). The top three pathways were clusters for *signal transduction*, *organelle biosynthesis*, and individual ps-PRS for *R anchoring of the basal body to the plasma membrane*.

### Secondary Analyses

In addition to WMH global-PRS, we included the six clusters and 55 individual ps-PRS that were enriched in at least one follow-up cohort in secondary analyses for effect modification by hypertension in UK Biobank, and for association with stroke or dementia in cohorts of older adults (**Figure 3**).

**Figure 3.**
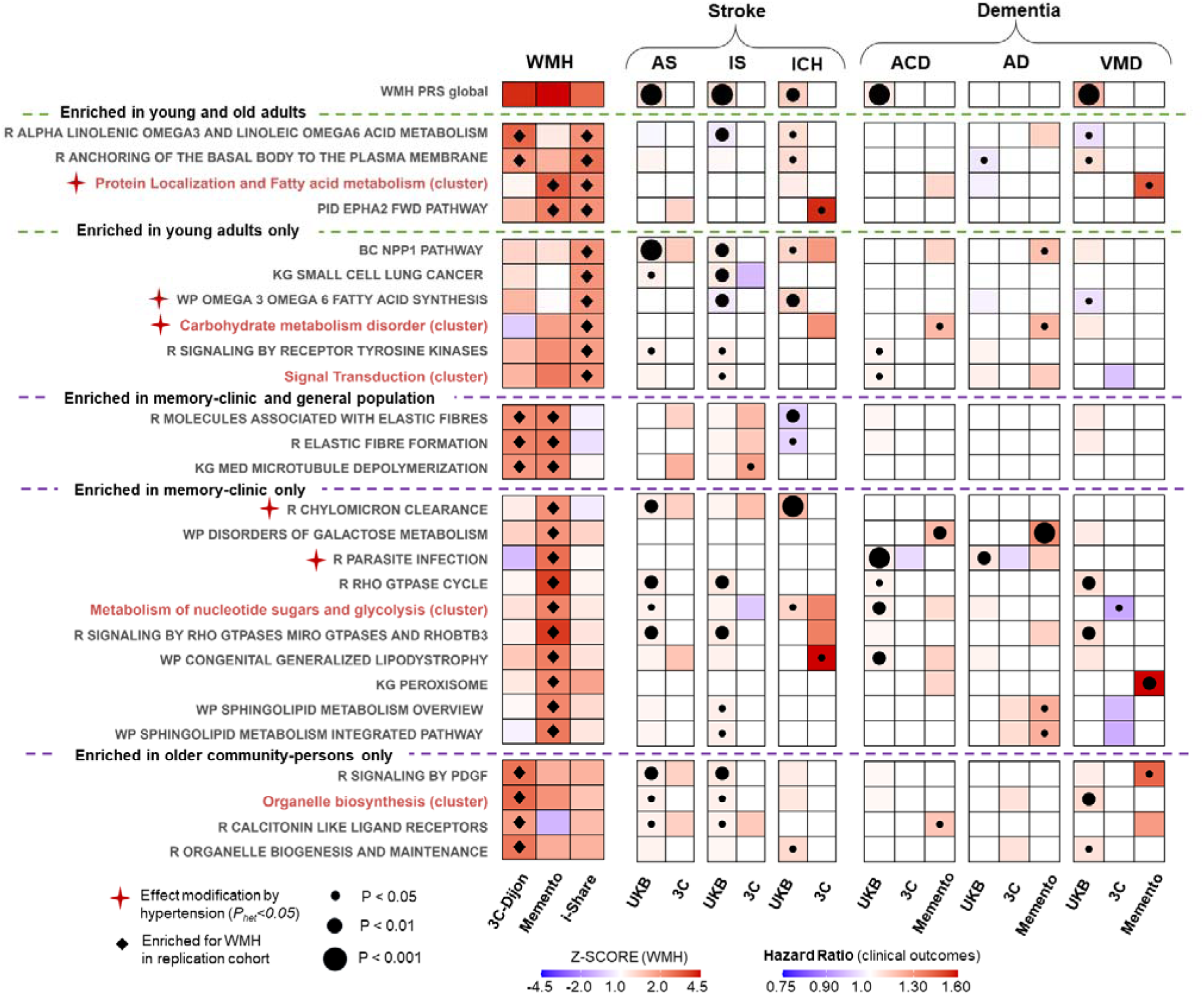
Prioritized findings from secondary analysis. We selected showing evidence of lifespan and clinical relevance—particularly, those enriched for WMH in young adults and showing association with stroke or dementia. Also shown are pathways enriched in older adults (memory-clinic or general population) showing stronger associations (*P<0.01*) or associated with multiple clinical outcomes. Red daggers indicate nominal evidence for effect modification by hypertension in association with WMH volume. AS = any stroke; IS = ischemic stroke; ICH = intracerebral hemorrhage; ACD = all-cause dementia; AD = Alzheimer’s Disease; VMD = vascular or mixed dementia; R = Reactome; PID = Pathway Interaction Database; WP = Wiki Pathways; KG = KEGG.

#### Effect modification by hypertension in the association of WMH (ps-)PRS with WMH volume

We investigated whether the association of WMH-PRS was modified by hypertension status in UK Biobank participants (*N= 12,520, of whom* 9,106 hypertensive and 3,414 non-hypertensive). The WMH global-PRS did not show notable differences in association between hypertension sub-groups, and most WMH ps-PRS showed consistent effect sizes as well (**Supplementary Fig. 3**, **Supplementary Table 15**). Seven WMH ps-PRS and clusters displayed nominally significant heterogeneity according to hypertension status (*P_het_<0.05*), each showing stronger effects in the hypertensive group: the *protein localization and import and fatty acid metabolism* cluster (*P_het_=0.028*) and two pathways within this cluster, *WP omega-3 and omega-6 fatty acid synthesis* (*P_het_=0.002*) and *R protein localization* (*P_het_=0.033*); the *carbohydrate metabolism (disorders)* cluster (*P_het_=0.024*), *R chylomicron clearance* (*P_het_=0.022*), and two pathways within the signal transduction cluster: *R FCG receptor dependent phagocytosis* (*P_het_=0.032*), and *R parasite infection* (*P_het_=0.032*) (**Figure 3**).

#### Association of WMH PRS with cSVD clinical complications, stroke and dementia

For incident stroke, study samples comprised 349,283 UK Biobank participants (9,212 incident stroke cases; 7,281 IS; 1,344 ICH), and 5,745 3C participants (232 incident stroke cases; 184 IS; 42 ICH). For incident dementia, we analyzed 355,180 UK Biobank participants (7,924 incident ACD cases; 2,728 AD; 1,187 VMD), 5,883 3C participants (662 incident ACD cases; 459 AD; 131 VMD), and 2,064 Memento patients (287 incident ACD cases; 197 AD; 37 VMD). Baseline characteristics of the samples used in these analyses are presented in **Supplementary Table 9**.

The WMH global-PRS was significantly associated with incident any stroke (HR_AS_=1.06 [1.038, 1.082], *P=2.6×10^-8^*) and its sub-types (HR_IS_=1.061 [1.037, 1.086], *P=1.6×10^-8^*; HR_ICH_ =1.083 [1.026, 1.143], *P=0.004*) in UK Biobank. For dementia, the WMH global-PRS was associated with incident ACD (HR_ACD_=1.051 [1.030, 1.080], *P=1×10^-5^*) and more strongly with VMD (HR_VMD_=1.168 [1.103, 1.237], *P=1.1×10^-7^*) in UK Biobank, but not with incident AD (HR_AD_=1.018 [0.98, 1.057], *P=0.35*). While the WMH global-PRS did not reach significance in association with any clinical outcomes in the much smaller 3C or Memento studies, associations were directionally consistent.

Of the 61 replicated WMH ps-PRS, 27 were nominally associated with incident stroke (*P<0.05*), and 32 with incident dementia outcomes in either population-based (UK Biobank or 3C) or memory-clinic (Memento) cohorts (**Figure 3, Supplementary Fig. 2**). Detailed results of all associations of WMH (ps-)PRS with clinical outcomes can be found in **Supplementary Table 10-14**.

Some interesting patterns are worth highlighting (**Figure 3**). Several cluster-level WMH ps-PRS showed nominal associations with incident stroke or dementia, the strongest associations (*P<0.01*) being observed between *organelle biosynthesis* and VMD or between *metabolism of nucleotide sugars and glycolysis* and ACD. At the individual pathway-level, several ps-PRS appeared to be associated (*P<0.01*) with incident IS specifically, namely *R signalling by PDGF*, *BC NPP1 pathway*, *KG small lung cancer*, and two Rho GTPase related pathways (*R Rho GTPase cycle* and *R signalling by Rho GTPases MIRO GTPases and RhoBTB3)*. The latter two were also associated (*P<0.01*) with VMD. Interestingly, the *BC NPP1 pathway* and *KG small lung cancer* pathways had replicated with WMH in young adults, suggesting lifespan effects. One ps-PRS, *WP disorders of galactose metabolism*, which had replicated in the memory-clinic setting (Memento) showed significant association with ACD (*P<0.01*) and AD (*P<0.001*). Another ps-PRS, *R chylomicron clearance*, showed robust association with ICH (*P<0.001*).

## Discussion

In this large multi-cohort study, leveraging global and ps-PRS for WMH derived from biobank-level data, we provide novel insights into the genetic underpinnings and biological pathways underlying WMH, the most-studied cSVD MRI-marker. We first confirmed that a global WMH PRS is associated with WMH volume in UK Biobank, with robust replication in three independent cohorts across the adult lifespan and clinical spectrum. Second, we identified 127 WMH ps-PRS enriched for WMH volume that clustered into ten coherent biological processes. These were validated across independent cohorts of young adults, community-dwelling older persons, and memory-clinic patients. Enriched WMH pathway clusters included notably those related to extracellular matrix and immune response, in line with previous findings in genomic and proteomic studies^12,13,56^, as well as new pathways such as lipid metabolism, ciliogenesis, and signal transduction. Several clusters and individual WMH ps-PRS showed distinct association patterns with incident stroke (ischemic or hemorrhagic) and dementia, particularly vascular and mixed subtypes.

There is emerging evidence that processes predisposing to cSVD take root early in life^3,13,15,17,57^. Here we highlighted several biological pathways that are enriched for WMH in both young and older adults with additional evidence for associations of several of them with stroke or dementia. These pathways converge on known cSVD hallmarks, including endothelial dysfunction, impaired blood–brain barrier function, altered fluid drainage, and vascular inflammation^58,59^.

Protein localization and fatty acid metabolism pathways emerged with the most robust evidence across the lifespan, both at the cluster and individual ps-PRS level, particularly related to omega-3 and omega-6 fatty acids. The *R alpha linolenic omega-3 and linoleic omega-6 acid metabolism* pathway was also associated with higher risk of incident ICH and lower risk of incident IS and VMD in UK Biobank, at nominal significance (P<0.05), suggesting possible utility in stratifying risk of cSVD complications, if validated independently. These findings align with metabolomic studies linking α-linolenic (omega-3) and linoleic (omega-6) fatty acid metabolites with WMH volume^60,61^. Omega-3 and omega-6 fatty acids are polyunsaturated fatty acids that come from the diet, and are involved in cell membrane structure, inflammation, as well as in vascular and brain health.^62^ Interestingly, hypertension appeared to modulate effects of these ps-PRS (at nominal significance), consistent with recent Mendelian randomization-based mediation analyses suggesting that hypertension is the strongest mediator of the potentially causal effect of hyperlipidemia on WMH^63^. Dysregulated lipid metabolism is central to low-grade systemic inflammation in metabolic syndrome and may underlie dysfunctional communication between the brain and the periphery found in these conditions^64^. Our findings thus reinforce lipid, and predominantly, fatty acid metabolism, as a possible central mechanism in cSVD across the lifespan.

Pathways related to organelle biosynthesis and maintenance, and specifically to ciliogenesis, also showed strong evidence across the lifespan, particularly those involved in basal body anchoring during primary cilia assembly. Several cilia-related and organelle biogenesis pathways were also enriched for WMH in older adults and associated with stroke and dementia. Overall, the organelle biosynthesis and maintenance (cilium) cluster was associated with VMD and, to a lesser extent, IS. Primary cilia are sensory organelles that mediate responses to shear stress from blood flow via nitric oxide and calcium channels in vascular endothelial cells ^65,66^. In neuronal and glial cells, they mediate signaling cascades for vascular-endothelial growth factor (VEGF), nitric oxide, and amyloid-β peptides. Dysfunction of these organelles can lead to aberrant fluid-sensing and contribute to hypertension and atherosclerosis^66–68^, and also disrupt vascular barriers (e.g., blood-brain-barrier)^69^. Interestingly, cilia proteins have been proposed as novel biomarkers of damaged endothelium^65^. Moreover, *TRIM47*, the putative causal gene at the most significant WMH risk locus (chr17q25)^13,70^, was recently shown to exert its effects by modulating the NRF2 pathway^71^, NRF2 being a transcription factor reported to control cilia formation and function^72^. Thus, our findings suggest that defects in primary cilia may play a central role in cSVD.

We provide evidence for signal transduction as a key driver of cSVD, demonstrating convergent signals at both the pathway-cluster level and within specific molecular cascades. Signal transduction encompasses the molecular networks that convert extracellular stimuli into coordinated cellular responses. At the cluster level, signal transduction was enriched for WMH in young adults (i-Share) and was associated with both IS and ACD. Beyond, we found specific ps-PRS associations involving VEGF, PDGF, ephrin (EPH), and EPHA2 forward signaling, all involved in processes relevant to cSVD. VEGF controls vascular permeability and angiogenesis^73^, PDGF regulates pericytes and blood-brain barrier integrity^74^, and EPH receptors are likely critical to development of tight junctions and blood-brain barrier permeability^75–77^. Of particular note, *PID EPHA2 forward signaling* was enriched for WMH across the lifespan and plasma EPHA2 and EPHB4 protein levels were recently found to be associated with WMH, cognition, stroke, and dementia^78^, with some drug repositioning perspectives^79^. Furthermore, VEGF and PDGF are key signaling pathways involved in aortic distensibility, itself causally linked to WMH^80^.

Extracellular matrix structure and function has previously been highlighted as a primary biological process underlying cSVD in pathways enrichment analyses from both genomic and proteomic studies^12^. Here, although the extracellular matrix pathway cluster did not replicate beyond UK Biobank, within this cluster we found robust evidence for the *BC NPP1* pathway, involving *ENPP1* and well-known cSVD genes *COL4A1/2*^12,81^. This pathway was enriched in young adults and uniquely associated with increased risk of incident IS and, at nominal significance, with incident ICH and AD. *ENPP1* regulates vascular calcification, a hallmark of coronary artery disease, through production of inorganic pyrophosphate^82^, as well as regulation of insulin receptor signaling and extracellular ATP levels.

In young adults, WMH may represent early stages of cSVD or early onset conditions like MS^83–86^. In this work, we showed that WMH global-PRS and 17 additional WMH ps-PRS were associated with WMH volume in young adults providing evidence that early-life WMH are consistent with white matter pathology found in older adults. In contrast, most ps-PRS enriched for WMH were distinct from those previously identified for MS^27^, in line with another genetic study showing that cSVD and MS largely do not have shared genetic underpinnings^83^.

In relation to clinical complications of cSVD, we originally demonstrate WMH global-PRS is predictive of stroke (both IS and ICH), ACD, and VMD, but not AD. Interestingly however, several WMH ps-PRS were associated with AD, including, at the cluster level, *immune response to viral infections* and *carbohydrate metabolism disorders*. Notably, infection-related pathways (parasite, viral) are associated with AD in population-based cohorts, supporting emerging evidence that they may uniquely contribute to the cSVD–AD link^87–89^.

We provide additional evidence that several other WMH ps-PRS may be of particular importance in older adults. *Elastic fiber* pathways, a key component of the arterial extracellular matrix^90^ and involved in atherosclerosis^91^, were enriched in older community-persons and memory-clinic patients. *R Chylomicron clearance* was significantly associated with ICH in UK Biobank, which is likely driven by the inclusion of *APOE* in this pathway^92^. Other notable pathways— *organelle biogenesis, RHO GTPases, PDGF signaling, sphingolipid metabolism*—showed consistent associations with both stroke and dementia, reinforcing shared cSVD mechanisms in these conditions.

In the memory clinic setting, several WMH ps-PRS related to *sphingolipid metabolism*, were specifically associated with risk of incident dementia. Indeed, sphingolipids form a core structural component of myelin, and their metabolites are implicated in neurodegeneration and neuroinflammation^93^. In particular, ceramides are highly dysregulated in stroke and cSVD patients and were shown to be associated with WMH volume and circulating biomarkers associated with brain injury^60,94–97^. The *WP disorders of galactose metabolism* pathway, significantly associated with AD, is also related to a subclass of sphingolipids—galactocerebrosides—that are essential for myelin structure and stability^98^. Sphingolipids are already an emerging target for diagnosis of treatment of cerebrovascular diseases, including cSVD and AD^99,100^. We also found the *KG peroxisomes* pathway to be associated with VMD in the memory-clinic. Peroxisomes play a critical role in neuroinflammation by regulating degradation of very-long-chain fatty acids and the metabolism of reactive oxygen species^101,102^. Particularly, peroxisome proliferator-activated receptor-γ (PPAR- γ), expressed in endothelial and vascular smooth muscle cells, is already used in treatment of diabetes and is being currently investigated as a therapeutic target against cSVD^103,104^. We propose that ps-PRS could have potential for prognostic enrichment in cSVD or AD clinical trials that target specific pathways, such as those involving sphingolipids or peroxisomes.

Our study’s primary strength is the comprehensive use of ps-PRS applied to cSVD MRI-markers. Unlike standard pathway enrichment analyses in GWAS (e.g., MAGMA), the ps-PRS approach directly quantifies an individual’s genetic susceptibility within specific biological pathways. This provides particular value for disentangling heterogeneous conditions such as cSVD and points towards which genetically driven processes may link WMH global-PRS to stroke and dementia. We also leverage genotyped and MRI data from multiple independent cohorts spanning the adult lifespan and clinical spectrum to robustly validate our findings and identify pathways implicated in cSVD across, or specific to, various life stages. We additionally highlight secondary analyses pertinent to understanding the link between WMH and hypertension, and downstream risk of stroke and dementia. Finally, our agnostic approach leverages existing pathway databases to comprehensively explore potential mechanisms without prior hypotheses—an ideal strategy for a heterogeneous disease like cSVD with largely unknown etiology. As more specific mechanisms emerge, hypothesis-driven approaches, as applied in AD^19,20,105^, will provide complementary insights.

Our study has several limitations. First, associations of ps-PRS are susceptible to confounding and pleiotropy, and some associations may reflect pleiotropic effects of genes within these pathways. Thus, ps-PRS findings should be interpreted with caution and ideally combined with complementary methods such as Mendelian randomization to answer etiological questions. Second, we acknowledge multiple testing burden with only few WMH ps-PRS being associated with clinical outcomes below stronger significance thresholds (*P<0.001)*. Thus, we position these analyses as exploratory, focusing on nominally consistent signals across follow-up cohorts and analyses. Third, while these cohorts together spanned a wide age range, they largely underrepresented midlife (ages 40–65) when vascular risk factors are most relevant, and follow-up cohorts (3C, Memento, i-Share) also had limited sample size. Additional limitations include variation in MRI protocols and WMH algorithms across studies, heterogeneous clinical outcome definitions and potential misclassification, particularly for algorithm-based stroke and dementia definitions in UK Biobank^106,107^. Finally, findings are limited to European ancestry, reducing generalizability^108^. As GWAS sample sizes increase in diverse populations, applying ps-PRS across ancestries will be critical to ensure equitable applications.

Overall, pathways enriched in younger adults may indicate early processes in cSVD development, whereas those found only in older or memory-clinic populations likely reflect later pathological stages predisposing to stroke or dementia. Pathways related to lipid metabolism, ciliogenesis, and signal transduction, newly described in this study, may underlie cSVD across the lifespan, highlighting the interplay between metabolic dysfunction, low-grade inflammation, and vascular risk factors such as hypertension and obesity. Our findings point to potential pathways to target therapeutically, including lipid subclasses (omega-3/6 fatty acids, sphingolipids, peroxisomes), NPP1 in the extracellular matrix, and tyrosine kinase signaling, particularly EPHA2. We also propose that primary cilia may act as key endothelial signaling hubs responsive to hemodynamic stress. Ps-PRS also have potential for risk stratifying cSVD complications, where certain biological endotypes may predispose more to cerebral amyloid angiopathy related risk of ICH, or risk of AD. Future studies should validate these mechanisms across ancestries and integrate multi-omics, especially metabolomics, to refine causal pathways. By capturing pathway-specific genetic susceptibility, ps-PRS can aid biomarker discovery, personalized prevention, and clinical trial stratification in cSVD.

In conclusion, in a multicohort setting we generated a robust global-PRS for WMH and highlighted pathway-specific PRS showing consistent associations across the lifespan and with cSVD clinical complications. These findings provide novel insights into cSVD mechanisms, with potential implications for therapeutic development and personalized prevention.

## Data availability

The data that support the findings of this study are available on request from the corresponding author. The data are not publicly available due to their containing information that could compromise the privacy of research participants.

## Acknowledgements

We would like to thank all UK Biobank, i-Share, Three-City and Memento participants, as well as the study teams (including the investigators and neurology teams) for their contributions and dedication to the study. We thank Dr. Murali Sargurupremraj for contributing the original WMH-GWAS summary statistics and providing access to these datasets. Computations were performed on the Bordeaux Bioinformatics Center (CBiB) and the CREDIM computer resources, University of Bordeaux. UK Biobank data access was granted through application number 94113.

## Funding

SD acknowledges support from the French National Research Agency and France 2030 (ANR-18-RHUS-0002, RHU-SHIVA; ANR-23-IAHU-0001, IHU-VBHI), Prix Burrus-FRM and NRJ-neurosciences, EU Horizon 2020 (grant No 754517). SD and QLG acknowledge support from the Fondation Recherche Alzheimer. TD was supported by the Digital Public Health Graduate Program (DPH), a PhD program supported by the French Investment for the Future Program (grant no. 17-EURE-0019).

The MEMENTO cohort is funded by the Fondation Plan Alzheimer (Alzheimer Plan 2008-2012), through the Plan Maladies Neurodégénératives (2014-2019), and the French Ministry of Research (MESRI, DGRI 2020-2024). This work was also supported by CIC 1401-EC, Bordeaux University Hospital (CHU Bordeaux, sponsor of the cohort), Inserm, and the University of Bordeaux. Genome-wide genotyping was funded by a grant (EADB) from the EU Joint Programme - Neurodegenerative Disease Research.

The Three-City (3C) Study is conducted under a partnership agreement among the Institut National de la Santé et de la Recherche Médicale (INSERM), the University of Bordeaux, and Sanofi-Aventis. The Fondation pour la Recherche Médicale funded the preparation and initiation of the study. The 3C Study is also supported by the Caisse Nationale Maladie des Travailleurs Salariés, Direction Générale de la Santé, Mutuelle Générale de l’Education Nationale (MGEN), Institut de la Longévité, Conseils Régionaux of Aquitaine and Bourgogne, Fondation de France, and Ministry of Research–INSERM Programme “Cohortes et collections de données biologiques.”

The i-Share study is conducted by the Universities of Bordeaux and Versailles Saint-Quentin-en-Yvelines (France). The i-Share study has received funding by the French National Agency (Agence Nationale de la Recherche, ANR), via the ‘Investissements d’Avenir’ program (grand number ANR-10-COHO-05) and from the University of Bordeaux Initiative of Excellence (IdEX). This project has also received funding from the European Research Council (ERC) under the European Union’s Horizon 2020 research and innovation program under grant agreement No 640643 and from the European Union’s Horizon 2020 research and innovation program under grant agreements No 643417, 667375 and 754517. This work was also supported by a grant overseen by the French National Research Agency (ANR) as part of the “Investment for the Future Programme” ANR-18-RHUS-0002 and as part of the France2030-funded precision and global vascular brain institute (IHU VBHI, ANR-23-IAHU-0001).

## Competing interests

The authors report no competing interests.

## Supplementary material

Supplementary material is available online.

## Notes

### Competing Interest Statement

The authors have declared no competing interest.

### Author Declarations

3C-Dijon: Ethics committee of the University Hospital of Kremlin-Bicetre gave ethical approval for this work. i-SHARE: The ethics committees of the Commission Nationale Informatique et Libertes (CNIL); CCTIRS (Comite Consultatif sur le Traitement de l'Information en matiere de Recherche dans le domaine de la Sante); CPP (Comite de Protection des Personnes); l'Agence Nationale de Securite du Medicament et Hors Produit de Sante (ANSM) gave ethical approval for this work. UK Biobank: The ethics committee of National Research Ethics Service Committee North West-Haydock (reference 48 11/NW/0382) gave ethical approval of this work. The MEMENTO cohort protocol has been approved by the local ethics committee (Comite de Protection des Personnes Sud-Ouest et Outre Mer III; approval number 2010-A01394-35)

