## Supplemental Material for "Pathway-specific polygenic risk scores reveal underlying mechanisms of cerebral small vessel disease"

**SUPPLEMENTARY MATERIAL**

### Supplementary Methods

#### MRI Sample exclusions

##### UK Biobank MRI

We excluded individuals who had an MRI prior to the reception date of the WMH GWAS manuscript from which we derived the risk variants and weights (January 26^th^, 2020) to avoid sample overlap bias^1^. We determined the UKBB-MRI sample on June 15^th^, 2025. As of this date, 16,320 individuals of European ancestry had available WMH volume recorded after January 26^th^, 2020. For related pairs (2^nd^ degree relatives, kinship > 0.0844), we randomly selected one individual to remove. We removed individuals with prevalent neurological conditions prior to MRI based on ICD-10 codes provided by field codes 41270 and 41280: Alzheimer’s Disease (ICD 10 codes G30-32), Parkinson’s disease (ICD-10 codes, G20), Stroke (I60-64), and multiple sclerosis (G35-37). We also additionally added prevalent cases for those with algorithmically-defined outcomes for stroke (field 42006), Parkinson’s disease (field 42032), and dementia (field 42018).

From the sample 16,360 we had the following exclusions:

- Missing Haplotype Reference Consortium (HRC) imputed genotyped data, n = 35
- In a related pair, n = 155
- Prevalent neurological conditions, n = 276
- Missing estimated total intracranial volume (eTIV), n = 574

Final sample: 15,320 participants

##### 3C-Dijon

1,924 participants in 3C-Dijon underwent MRI.

Exclusions:

- Low-quality MRI, n = 10
- No WMH and white matter volume mask, n = 113
- No viable genetic data, n = 168
- Prevalent stroke or dementia or no data on stroke or dementia history, n = 106
- Brain tumor, n = 6
- 3^rd^ degree relatedness, n = 78

Final Sample: 1,443 participants

##### i-Share

For the present study, the population comprised i-Share participants taking part in both MRi-Share and bio-Share and for whom brain MRI and genome-wide genotype data passed quality control filters. Of 1,954 participants who took part in MRi-Share, 1,856 had a usable (after quality control, QC) brain MRI. Genome-wide genotype data were available for 1,862 individuals. In total, 1,784 participants had both high-quality brain MRI and genome-wide genotype data available.

Additionally:

- Seven individuals were excluded with a diagnosis or suspicion of multiple sclerosis or radiologically isolated syndrome
- 151 were excluded due to non-European ancestry
- 48 were excluded due to 3^rd^ degree relatedness

The final i-Share sample had 1,578 eligible participants (mean age=22.1±2.3 years, 71.7% female).

##### Memento

The Memento study is a memory clinic-based cohort aimed to investigate the natural history of AD and related disorders in a large group of participants with MCI or subjective cognitive complaints. Memento participants were recruited from 26 French memory clinics in 2011-2014, with a follow-up every 6-12 months over 5 years^2^. We excluded those with missing WMH volume or genetic data, with prevalent stroke, dementia or Parkinson’s disease, or a 3^rd^ degree relative in the sample, resulting in 1,831 eligible participants (mean-age=71.1±8.5 years, 62.2% female).

2,246 individuals underwent MRI

Exclusions:

- Missing WMH volume, n = 142
- Extreme intracranial volume, n = 1
- No viable genetic data, n = 170
- Prevalent dementia, n = 18
- Prevalent stroke, n = 79
- Parkinson’s disease, n = 4
- Relatedness, n = 1

Final sample : 1,831 participants

#### MRI Acquisitions and WMH calculation

##### UK Biobank

Details of the UK Biobank MRI protocol and documentation is publically available (<https://www.fmrib.ox.ac.uk/ukbiobank/>) and image-derived phenotyps are fully described in a previous publication^3^. In brief, the data were acquired with a standard Siemens Skyra 3T 32-channel head coil. Total volume of WMH was estimated via T_1_-weighted MPRAGE and T_2_-weighted FLAIR volumes. The total volume of WMH calculated using T_1_-weighted MPRAGE and T_2_-weighted FLAIR volumes segmented by the BIANCA tool^4^.

##### 3C-Dijon

MRI scans in 3C-Dijon were acquired from a 1.5-Tesla Magnetom scanner (Siemens, Erlangen, Germany)^5^. T1- and T2-weighted images of each subject were aligned to each other using the AIR package. These images were analyzed with the optimized Voxel-Based Morphometry (VBM) protocol, using Statistical Parametric Mapping 99 (SPM99) that were modified in order to take into account the structural characteristics of the aged brain. Fully automated image processing software was developed to detect, measure, and localize white matter hyperintensities (WMH)^6^.

##### i-Share

1,832 i-Share participants participated in the MRi-Share sub-study and underwent MRI acquisition protocol designed to closely emulate that which was done in UK Biobank^7^. T1-weighted and T2-weighted FLAIR structural scans were acquired with a Siemens 3 Tesla Prisma scanner with a 64-channels head coil between November 2015 and November 2017. Due to low WMH volume in young adults, WMH volume was derived with the SHIVA-WMH tool developed to improve detection of small and sparse WMH^8^.

##### Memento

In Memento, 86% of participants had a 3 Tesla or 1.5 Tesla MRI scan at their respective memory clinics^2^. Images were harmonized by the Center for Automated Treatment of Images (CATI)^9^, and standard acquisitions were applied according to a systematic qualification procedure ensuring parameter uniformity and image quality. MRI sequences for T1-weighted and T2-weighted FLAIR were conducted to ensure compatibility with Alzheimer’s Disease Neuroimaging Initiative (ADNI) protocol. WMH volume was measured with White Matter Hyperintensities Automated Segmentation Algorithm software^10^ and quantitatively verified by two trained physicians.

#### Genotyping collection and quality control

##### UK Biobank

Quality control and imputation with the Haplotype Reference Consortium reference panel are fully described elsewhere^11^. We were granted access to UK Biobank data through application number 94113. We used previously generated relatedness variables available of the UK Biobank Research Analysis Platform to identify related pairs (kinship ≥ 0.0844) and randomly selected individuals in each pair for exclusion (n=155).

### 3C

For 3C participants, genome-wide genotyping was performed at the Centre National de Génotypage in Evry (France) using the Illumina Human610Iquad BeadChips. Standard quality control procedures were initially conducted on raw genotyped data. This included removing SNPs due to missingness (<98% call rate), low minor allele frequency (MAF) (<1%), non-autosomal location, and deviation from Hardy-Weinberg equilibrium (p<0.001). Participants were removed with discordant sex information, excessive missingness (<95% call rate), high heterozygosity (±3SD), or divergent ancestry (±6SD)^12^. Additionally, sample pairs were identified with 3^rd^-degree relatedness with KING software (kinship > 0.0625), and were removed. The raw genotyped data was then imputed using the HRC reference panel^13^.

##### i-Share

Genome-wide genotyping of 1 872 i-Share participants was performed using the Affymetrix Precision Medicine Axiom Array at McGill Genome Center (Canada). After quality control, genotype data were available for 1 862 participants (7 participants were removed due to sex discrepancies, 2 participants who appeared to be duplicates but not twins, and one participant with a kinship coefficient >0.0625 (third degree related) with more than 20 other participants, suggesting a possible sample contamination (KING software) ^14^. After applying standard quality control procedures (SNP call rate <98%, Hardy-Weinberg Equilibrium p<0.001), we imputed the genotypes on the HRC reference panel^15^.

##### Memento

Pre-imputation QC included removal of SNPs with MAF<0.01, callrate<0.98 and HWE<0.001, removal of samples with callrate <0.05, heterozygosity beyond 3SD, failed sex-check using genotype data of X-chromosome, related sample based on IBD (pi_hat>0.1875), PCA outliers beyond 6SD of PC1 and PC2. The raw genotyped data was then imputed using the HRC reference panel^13^. Additional related pairs were identified with 3^rd^-degree relatedness with KING software (kinship > 0.0625). We randomly removed one individual from each related pair.

#### PRS Methods

In clumping and thresholding, SNPs are first clumped to account for LD, selecting the most significant within ±250kb, and removing any variants correlated with the index SNP (r^2^>0.1). A range of p-value thresholds (p_t_=5x10^-8^, 1x10^-6^, 1x10^-4^, 0.01, 0.1, 0.2, 0.3, 0.4, 0.5, 1) is then applied to shrink effect sizes for SNPs with a p-value over the threshold to zero. Alternatively, PRS-CS is a Bayesian approach that utilizes a continuous shrinkage prior on SNP effect sizes to directly model LD and limit loss of information^16^. PRS-CS requires tuning of the global shrinkage parameter (ϕ) representing prior knowledge about the sparseness of the underlying genetic architecture. The starting ϕ value can be manually set or tuned ‘automatically’, thus, we used φ=1, 1x10^-2^, 1x10^-4^, 1x10^-6^, 1x10^-8^, or ‘auto’.

#### PRSet Enrichment

The 3,794 curated biological pathways in the Canonical Pathway in the Molecular Signatures Database (MSigDB v2023.2) includes pathways from the Kyoto Encyclopedia of Genes and Genomes (KEGG)^17^, REACTOME^18^, BioCarta^19^, wikiPathways^20^, and the Pathway Interaction Database (PID) ^21^.

PRSet relies on empirical, “competitive” P-values derived from a permutation-based procedure to determine pathway enrichment. PRSet is applied on clumped SNPs (r^2^<0.1) without any p-value thresholding. This is to ensure that all genes in the pathway are represented regardless of their underlying association to the trait. The permutation-based procedure also accounts for the number of SNPs included in a given pathway, so as not to privilege pathway enrichment towards pathways with a larger number of genes.

In brief, the method is summarized in the PRSet paper:

1. A “background” pathway containing all genic SNPs is constructed, and clumping is performed within this pathway. For pathways with m SNPs, N null pathways are generated by randomly selecting m “independent” SNPs from the “background” pathway.
2. The competitive P-value can then be calculated as

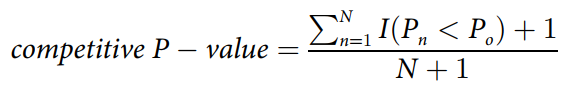

Where: I(.) is an indicator function, taking a value of 1 if the association P-value of the observed gene set (P0) is larger than the one obtained from the nth null set (Pn), and 0 otherwise. A pseudo-count of 1 is added to the numerator and denominator to avoid competitive P-values of 0 and conservatively counting the observed gene set as 1 potential null set [54]. One consideration of this permutation procedure is that the smallest achievable competitive P-value is 1/(N+1), which can lead to difficulties in ranking highly significant gene sets.

This method is recommended by the authors over standard regression procedures and multiple testing correction.

#### Clinical Outcomes Secondary Analysis

Below you will find study specific exclusions for the clinical outcome secondary analysis, and outcome definitions. Each study and outcome also required specific modeling in order to fit cox proportional hazard models that satisfied the cox proportional hazard assumption.

##### UKBB

For analysis of clinical outcomes in UK Biobank, we excluded participants with brain MRI to avoid overlap with the WMH GWAS and also with the UKBB-MRI sample used to identify enriched WMH ps-PRS. We also exclude those of non-European ancestry, and those in a related pair (2^nd^ degree relative). For analyses of stroke or dementia outcomes, we exclude individuals with prevalent conditions. We also remove individuals with self-reported stroke. Individuals were censored at clinical outcome, death, or end-of-follow-up was considered as of May 31, 2022 – the latest date where hospital records are mostly complete from Wales, Scotland, and England. We ascertained stroke and dementia outcomes using algorithmically-defined stroke and dementia outcomes to identify incident stroke cases defined during follow-up with UK Biobank fields 42006-42010 (any stroke, IS, and ICH), and 42018-42022 (dementia, AD, and vascular dementia) (<https://biobank.ndph.ox.ac.uk/showcase/ukb/docs/alg_outcome_main.pdf>).

###### Exclusions:

- Non-European ancestry, n = 93,579
- Relatedness, n = 3,535
- Brain MRI (as of June 15^th^, 2025), n = 49,810

Stroke-specific exclusions:

- Prevalent stroke, n = 6,046
- Self-reported stroke, n = 28

Sample size for incident stroke analyses: N=349,283

- Undefined stroke cases removed from subtype-specific analysis: n = 587

Dementia-specific exclusions:

- Prevalent dementia, n = 176
- Self-reported incident dementia, n = 1

Sample size for incident dementia analyses: N= 355,180

- Undefined dementia cases removed from subtype-specific analysis: n = 4,009

End-of-follow-up was considered as of May 31, 2022 – the latest date where hospital records are mostly complete from Wales, Scotland, and England.

###### Stroke Modeling in UKBB

- **Any Stroke:** cox proportional hazard models were stratified by sex and geno-chip in order to satisfy proportional hazard assumptions, allowing baseline risks to vary across these strata. Models were further adjusted by 10 genetic principal components.
- **ICH or IS:** cox proportional hazard models were stratified by sex. Models were adjusted for genotype chip and 10 principal components.

###### Dementia Modeling in UKBB

- **Any Dementia:** cox proportional hazard models were stratified by sex, and adjusted for genotype chip, 10 PCs, and number of APOE-ε2 and APOE-ε4 alleles (0, 1 or 2)
- **AD:** cox proportional hazard models were adjusted for sex, genotype chip, 10 PCs, and number of APOE-ε2 and APOE-ε4 alleles (0, 1 or 2)
- **VMD:** cox proportional hazard models were stratified by sex, and adjusted for genotype chip, 10 PCs, and number of APOE-ε2 and APOE-ε4 alleles (0, 1 or 2)

##### Three Cities (3C)

In 3C, an expert panel reviewed potential stroke cases identified at each follow-up visit with participants or informants for deceased participants^22^. Expert panels of neurologists reviewed all available clinical information, confirming diagnosis of stroke and its subtype (ischemic or haemorrhagic) according to the International Classification of Diseases – 10^th^ Edition^23^. Diagnosis of dementia followed a three-step procedure at baseline and at each follow-up visit^24^. First, all participants underwent a battery of neuropsychological tests at baseline and each follow-up visit by a trained psychologist. Second, a neurologist examined all the participants in Montpellier and in Bordeaux at baseline. In Dijon, participants screening positive for dementia based on their neuropsychological performance underwent further clinical examination. At each follow-up, all participants with suspected dementia were examined by a neurologist to establish provisional diagnosis. We combined vascular and mixed dementia diagnoses to capture cases with a more substantial vascular component.

Individuals were censored at death, clinical outcome, or end of follow-up. If an individual died 2.5 years after their last visit, they were censored at the date of their last visit since it is unknown if they had a stroke or developed dementia during this period.

###### Exclusions:

Full sample: 9,294

***Stroke***

- No viable imputed genotypes, n = 2,805
- Missing stroke information, n = 294 (could be due to just insufficient follow-up)
- Relatedness, 3^rd^ degree, n = 177
- Prevalent stroke, n = 273

**N = 5,745**

***Dementia***

- No viable imputed genotypes, n = 2,805
- Relatedness, 3^rd^ degree, n = 181
- Prevalent dementia, n = 121
- Insufficient follow-up, n = 304

N = 5,883

###### Stroke Modeling in 3C

- **Any Stroke:** cox proportional hazard models adjusted for sex, study centre, and 4 genetic principal components
- **ICH:** cox proportional hazard models were stratified by sex, and adjusted for study centre, and 4 PCs
- **IS:** cox proportional hazard models adjusted for sex, study centre, and 4 genetic principal components

###### Dementia Modeling in 3C

- Cox proportional hazard models for dementia outcomes were all stratified by study centre, and adjusted for sex, 4 PCs, and dosage of APOE-ε2 and APOE-ε4

##### Memento

In Memento, trained neurologists administered a battery of neuropsychological tests at baseline, and assessed dementia status at each follow-up visit^2^. In both cohorts, an independent panel of expert neurologists reviewed and validated all possible dementia cases according to the *Diagnostic and Statistical Manual of Mental Disorders* (Fourth Revision) criteria. We combined vascular and mixed dementia diagnoses to capture cases with a more substantial vascular component.

Individuals were censored at death, dementia, or end of follow-up. If an individual died one year after their last visit, they were censored at the date of their last visit since it is unknown if they developed dementia during this period.

###### Exclusions

Full sample: 2,323

- Missing phenotype information, n = 119
- No viable imputed genotypes, n = 87
- Prevalent dementia, n = 19
- Insufficient follow-up, n = 34

N = 2,064

###### Study-specific Modeling in Memento

In Memento we compared random effect vs. fixed effects for site ID since there were 33 of them, in order to control the number of parameters in the model. We compared both modelling approaches using AIC, for which mixed effects consistently had better fit. For Memento we used mixed effect cox proportional hazard models^25^ using the coxme R package^26^ allowing for a random effect based on memory-clinic site, of which were there were 33. Models were further adjusted for sex, 7 PCs, and dosage of APOE-ε2 and APOE-ε4.

#### Hypertension definition in UKBB

Where available, we used automated DBP and SBP readings at the imaging assessment (field codes 4079 and 4080, instance 2) to ascertain blood pressure at baseline. We also used self-reported medical conditions data at the imaging assessment to ascertain whether an individual was taking blood pressure medication – field codes 6153 and 6177 for females and males respectively. We classified individuals as hypertensive if they had an automated SBP ≥ 140 mmHg or DBP ≥ 90 mmHg or if they self-reported blood pressure medication. Individuals were excluded if they did not have blood pressure readings and if they also did not report taking blood pressure medication or did not self-report any medications. A total of 2,800 individuals were excluded from this analysis.

In total, 4,306 self-reported taking blood pressure medication at the time of the imaging assessment, 7,469 had SBP ≥ 140 mmHg or DBP ≥ 90 mmHg.

### Supplementary Figures

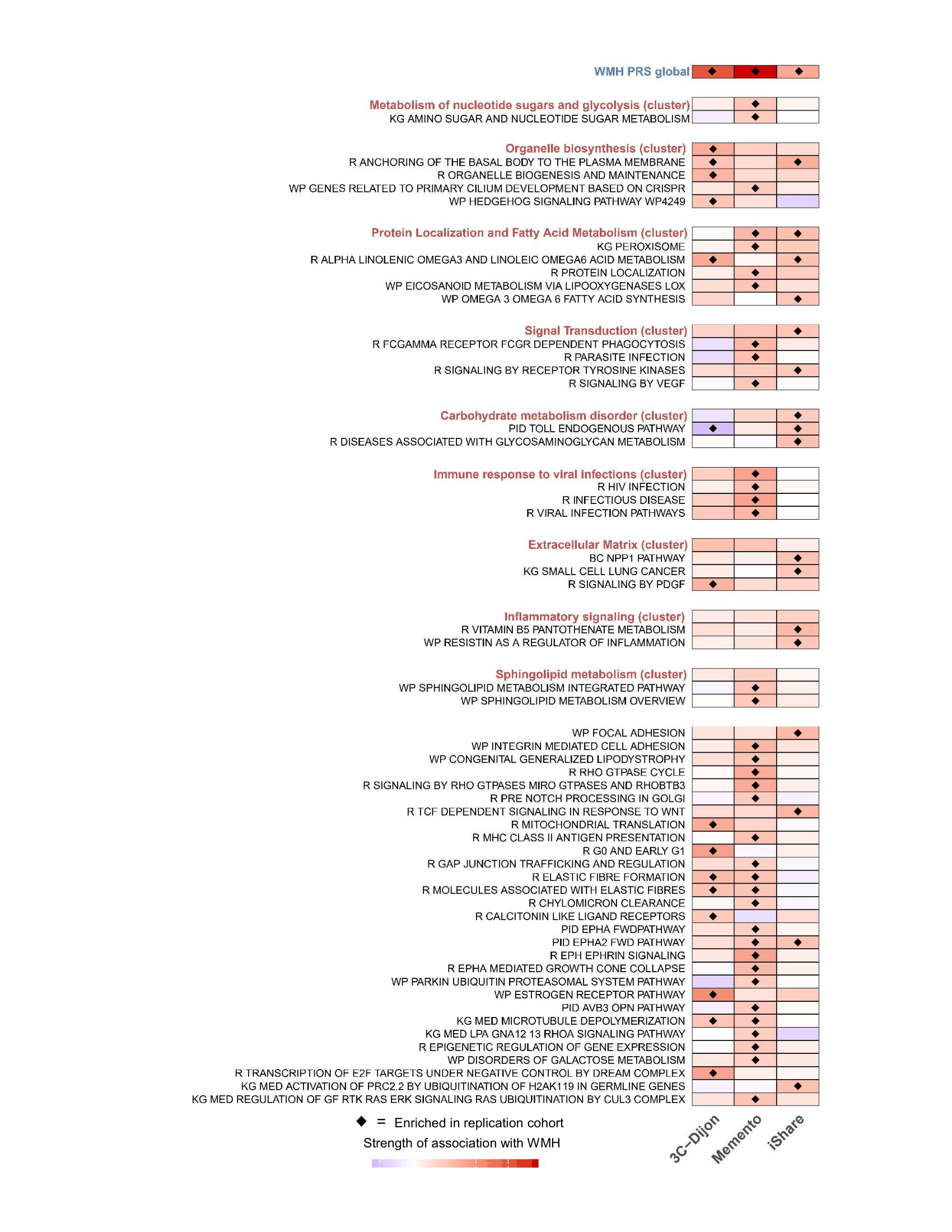
**Supplementary Figure 1.** Enrichment of WMH ps-PRS in Follow-up Cohorts. Pathways are grouped based on their biological cluster, represented by the first pathway in each grouping indicating cluster-level enrichment. Only individual-level pathways that are enriched in at least one follow-up cohort are shown. The last, larger group are all pathways that did not belong to any cluster.

##### Supplementary Figure 2. Associations of all WMH-psPRS included in the secondary analysis in Cox proportional hazard models. All models are adjusted for adjusted for sex, genetic PCs and other study-specific covariates. Dementia was additionally adjusted by APOE-ε2 and ε4 dosage.

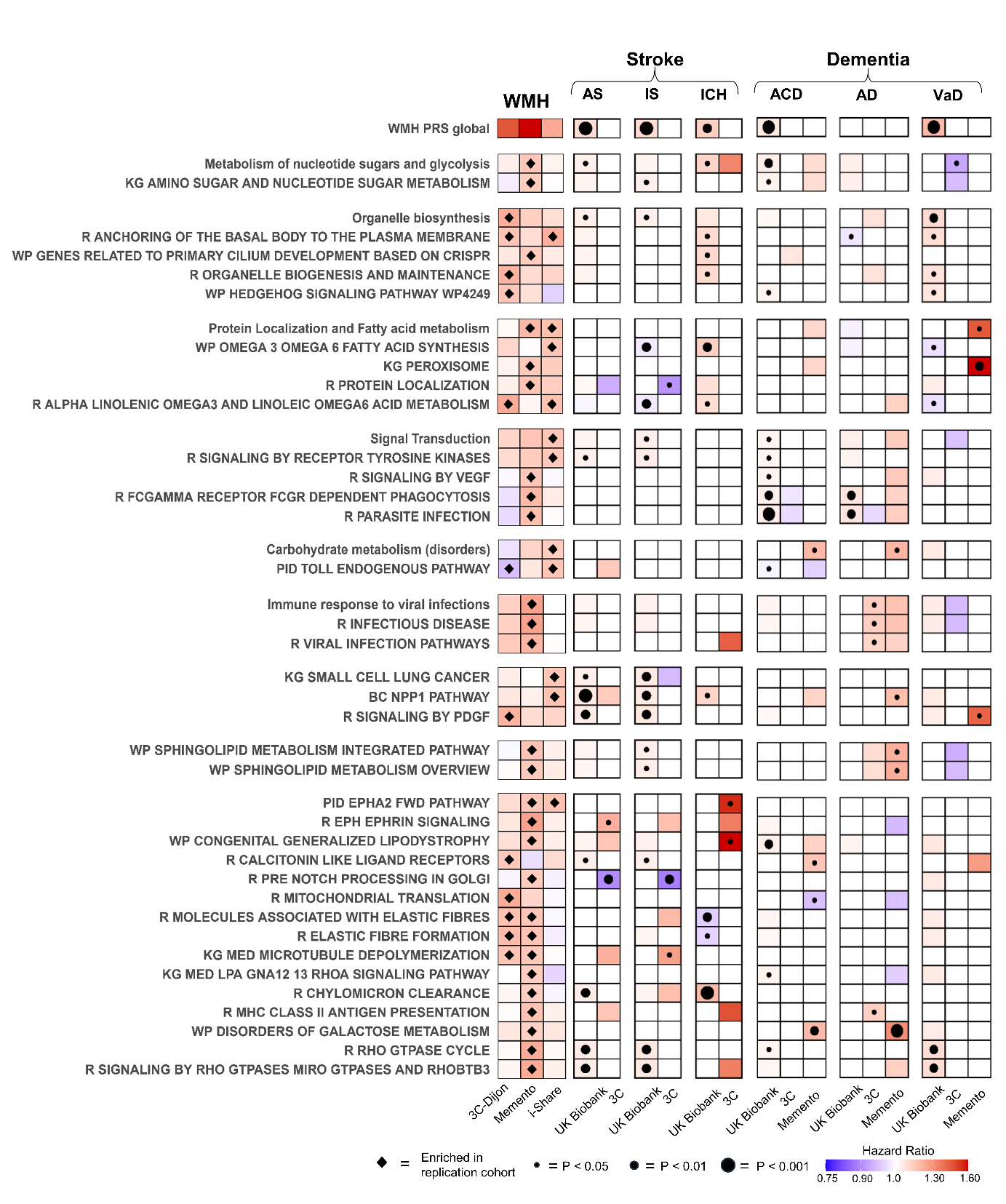

##### Supplementary Figure 3. Effect modification by hypertension in the association of WMH-(ps)PRS with WMH volume. Linear regression models for WMH volume were adjusted baseline covariates: age at MRI, sex, 10PCs, genotyping chip and total intracranial volume. We show WMH ps-PRS with a heterogeneity P-value < 0.3. * indicates nominal significance (p<0.05

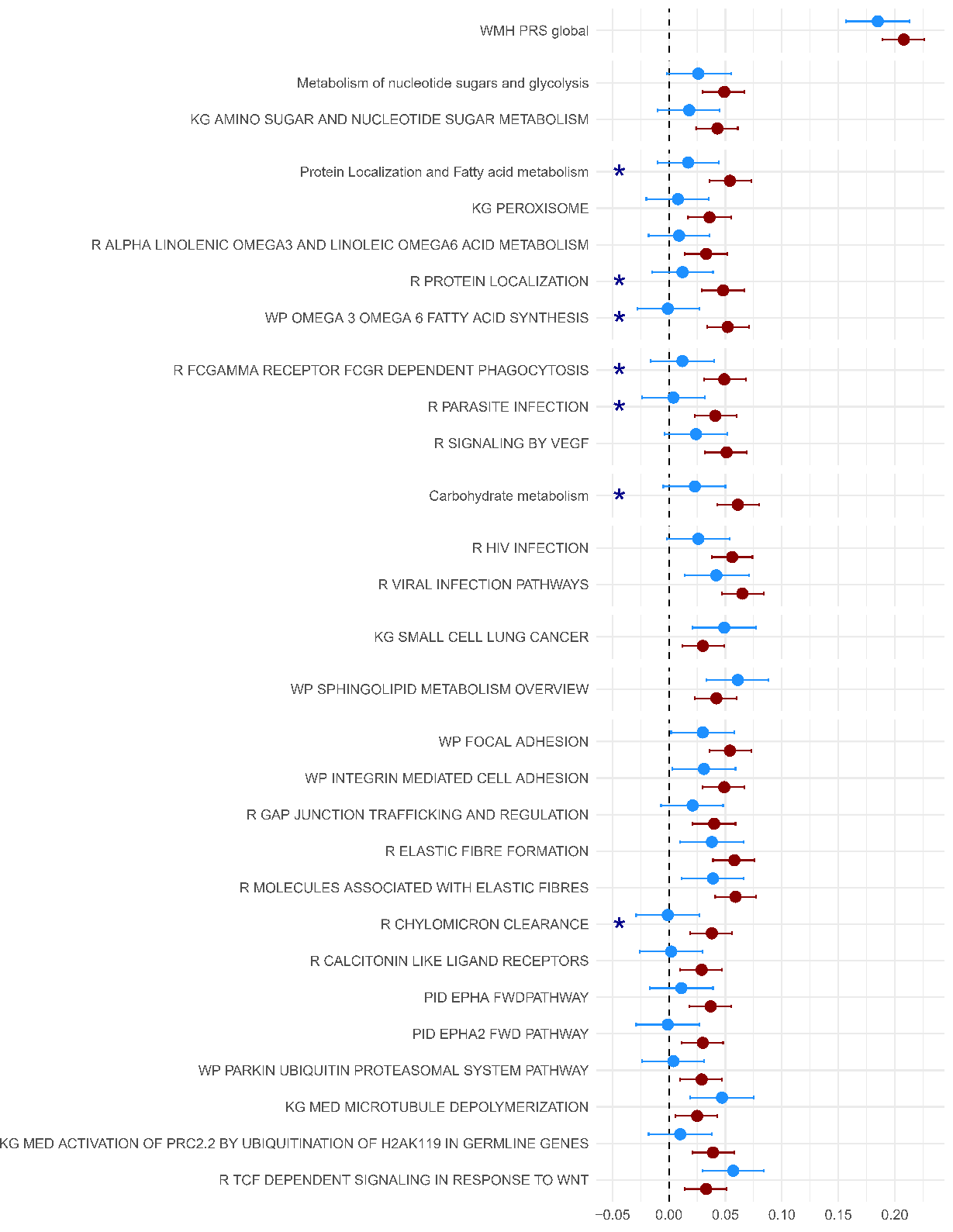
